# The frameshifted VNTR length determines the kidney survival in *ADTKD-MUC1*

**DOI:** 10.64898/2026.10.28.26365901

**Authors:** Mouad Ouahmane, Hassan Saei, Aurélien Caux, Jessica Kachmar, Zaina Ait Arkoub, Maxime Fercocq, Manon Mautret-Godefroy, Olivier Gribouval, Vincent Morinere, Bertrand Knebelmann, Laurence Heidet, Corinne Antignac, Collaborators, Guillaume Dorval

**Affiliations:** Université Paris Cité. Department of Genomic Medicine for Rare Diseases, Necker–Enfants Malades Hospital, Assistance publique, Hôpitaux de Paris (AP-HP), Paris, France; Laboratory of Hereditary Kidney Diseases, Inserm UMR 1163, Institut Imagine, Université Paris Cité, Paris, France; Centre Hospitalier de l’Université de Montréal, Department of Nephrology, Montreal, Quebec, Canada; Université Paris Cité. Department of Adult Nephrology, Necker–Enfants Malades Hospital, Assistance publique, Hôpitaux de Paris (AP-HP), Paris, France; Université Paris Cité. Department of Pediatric Nephrology, Necker–Enfants Malades Hospital, Assistance publique, Hôpitaux de Paris (AP-HP), Paris, France; Reference Center for Inherited Kidney Diseases in Adult and Children (MARHEA)

**Keywords:** *ADTKD-MUC1*, genotype/phenotype correlation, Long-Read Sequencing Technology, genetics, kidney genetics

## Abstract

**Introduction:** Autosomal dominant tubulointerstitial kidney disease due to *MUC1* variants (ADTKD-*MUC1*) is caused by pathogenic variants leading to a frameshift in the *MUC1* gene, most often because of the duplication of a cytosine in a 7-cytosine stretch within the coding variable number tandem repeat (VNTR). It has been regarded as genotypically uniform despite marked clinical heterogeneity. However, as the duplication can arise in any repeat unit, the length of the toxic MUC1-frameshift protein (MUC1-fs) may differ substantially between patients. We thus hypothesised that MUC1-fs length may determine disease severity.

**Methods:** In this retrospective cohort study, we evaluated 49 patients with genetically confirmed ADTKD-*MUC1*. Mutant VNTR alleles were reconstructed by Oxford Nanopore long-read sequencing. The correlation between total and MUC1-fs length and age at kidney failure (KF) was assessed using statistical models accounting for familial clustering.

**Results:** Median KF-free survival was 43 years (95% CI 37–55) for MUC1-fs above the median (>780 amino acids) versus 65 years (95% CI 54–not reached) below it (log-rank p = 0.0039). MUC1-fs length was associated with KF (HR 1.19 per 100 amino acids, 95% CI 1.03–1.37, p = 0.015). The estimate was stable across five handlings of familial clustering, including leave-one-family-out and random one-per-family sampling. Longer MUC1-fs was also correlated with faster eGFR decline. By contrast, the total length of the mutant protein showed a weaker association with KF, and no association when both measures were modelled together, suggesting that the signal lies in the frameshifted portion rather than in overall allele size.

**Conclusions:** In this exploratory cohort, MUC1-fs length, determined by the position of the duplication within the VNTR, was associated in a graded manner with age at KF, independently of familial clustering. These findings suggest that ADTKD-*MUC1* is not genotypically uniform and that reporting the frameshift position could refine prognosis.

**Lay Summary:** ADTKD-*MUC1* is an inherited kidney disease. Nearly all patients carry the same mutation, which has supported the view that the disease runs a similar course in everyone. It does not. The mutation falls within a long tract of repeated DNA, and its position along that tract varies between families. That position sets the size of the abnormal protein the kidney cells accumulate: the earlier the mutation, the larger the abnormal protein. Routine genetic tests cannot read this tract reliably, so we used sequencing able to span it from end to end. Across 49 patients from 24 unrelated kindreds, median age at kidney failure was 43 years in the group making the largest abnormal protein and 65 years in the group making the smallest. Determining where the mutation falls therefore adds prognostic information that the standard test cannot give, and larger cohorts should now confirm it.

## Introduction

Autosomal dominant tubulointerstitial kidney disease (ADTKD) due to *MUC1* variants is one of the main forms of ADTKD yet remains widely underdiagnosed. Since the disease is often clinically silent (bland urinary sediment and minimal or absent proteinuria), it is frequently detected only at an advanced stage^1,2^. Dominant frameshift variants generate a truncated neoprotein, MUC1-frameshift (MUC1-fs), which accumulates in a TMED9-positive early-secretory compartment of renal tubular cells, causing cellular stress, inflammation and tubulointerstitial damage. The neoprotein itself is the driver of the disease and a potential therapeutic target^3,4^.

The clinical course of ADTKD-*MUC1* is variable. Median age at kidney failure (KF) was 36 years (IQR 30–46) in the largest genetically confirmed cohort, with onset ranging from the second to ninth decade and variability between and within families^5^. Because the genotype appears uniform, this variability has been attributed to modifiers^5^. Most pathogenic variants are single-cytosine duplications within a seven-cytosine tract, most commonly NM_001204287.2:c.428dup (also designated 59dupC or 27dupC), predicted to generate the same frameshift and the same premature stop codon^3^. Genotype-derived prognostic information is therefore currently lacking to guide counselling or transplant planning.

However, this assumption overlooks the architecture of the locus. The *MUC1* coding variable number tandem repeat (VNTR) comprises 20 to 125 units of a 60-mer motif, each encoding a 20– amino acid block^3,6,7^. The duplication can arise in any unit: its position determines how many repeats are translated in frame and hence the length of MUC1-fs, which may differ by more than twofold between patients carrying the same reported variant. Resolving that position requires contiguous sequencing across the GC-rich VNTR, feasible only with long-read technologies^3,6,7^.

We thus hypothesized that MUC1-fs may play a role in disease severity. To assess this, we evaluated whether there was a correlation between MUC1-fs length and age at KF as well as eGFR decline, accounting for familial non-independence.

## Short methods

We retrospectively evaluated 49 patients with genetically confirmed ADTKD-*MUC1* from 24 unrelated kindreds, in whom mutant *MUC1* VNTR alleles were reconstructed by Oxford Nanopore long-read sequencing using VNTRpipeline^8^. We then analysed the association between genotypic features of the mutant allele (VNTR repeat number, position of the mutated repeat, and resulting total and MUC1-fs lengths) and disease severity (age at KF and rate of eGFR decline). Each kindred, including singleton kindreds, was treated as one independent unit, and age was used as the time scale. Full methods are provided in the supplementary material.

## Results

### Cohort characteristics

Forty-nine patients carrying a *MUC1* 59dupC variant, identified with VNTyper^7^, were included. They comprised 36 affected individuals from 11 families and 13 singleton kindreds, giving 24 independent units for clustered analyses. Thirty-three patients (67%) had reached KF at the time of data collection, at a median age of 49 years (95% CI 44–67). Cohort characteristics are summarised in **Supplementary Table S1**.

The *MUC1*-VNTR was reconstructed from ONT sequencing of targeted amplicons and analysed with VNTRpipeline^8^. For 5 patients, we performed biological replicates to ensure reproducibility with a concordant VNTR structures. Repeat counts were converted into protein length in amino acids (aa): 20 amino acids per repeat unit. The median total protein length was 1560 aa and the median MUC1-fs length 780 aa (**Supplementary Table S1**).

### Genotype and kidney survival correlation

Patients were dichotomised at the median value of each measure (total protein length and MUC1-fs length). KF-free survival differed significantly depending on MUC1-fs length: median survival was 43 years (95% CI 37–55) in patients above the median (> 780 aa), versus 65 years (95% CI 54– not reached) in those below it (log-rank p = 0.0039; **Figure 1A**). Stratification by total protein length (1560 aa) showed a similar but less pronounced difference, with median survival of 43 years (95% CI 39–not reached) versus 55 years (95% CI 54–not reached; log-rank p = 0.026; **Figure 1B**).

**Figure 1.**
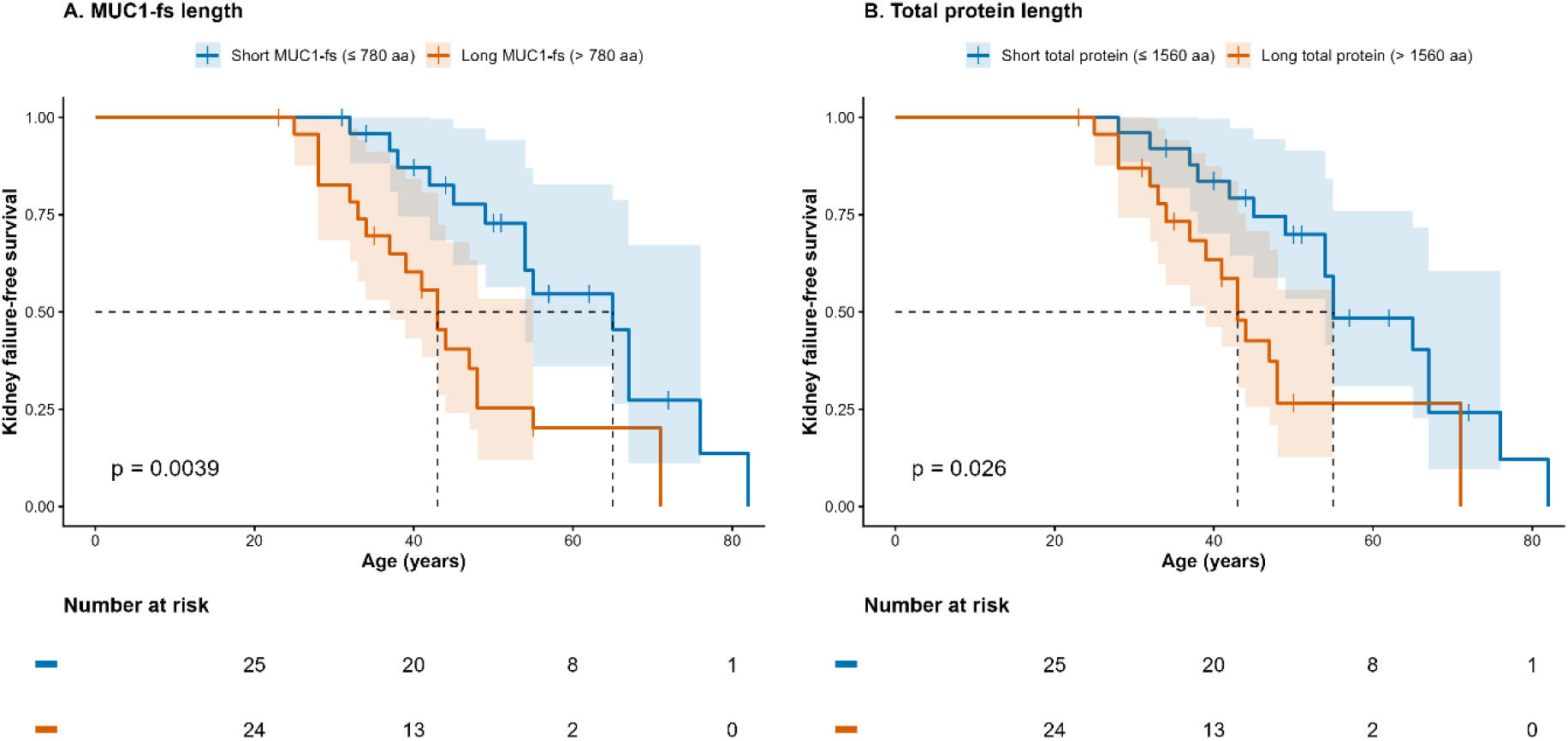
Kidney failure-free survival according to total and MUC1-fs protein length. Kaplan-Meier estimates of kidney failure-free survival, with age as the time scale, in 49 patients with ADTKD-*MUC1*. Patients were dichotomised at the median value of MUC1-fs length (**A**, 780 aa) and of total protein length (**B**, 1560 aa), each yielding groups of 25 and 24 patients. Shaded areas represent 95% confidence intervals and vertical ticks denote censored observations. Dashed lines indicate the median kidney failure-free survival of each group. P-values are from the log-rank test. Numbers at risk are shown below each panel, colour coded as in the curves. (**A**) Median kidney failure-free survival was 43 years (95% CI 37–55) for long versus 65 years (54–NR) for short MUC1-fs. (**B**) The corresponding values for total protein length were 43 years (39–NR) and 55 years (54–NR). NR, not reached.

Continuous Cox models with robust standard errors clustered on family were used for inference (**Supplementary Table S2**). MUC1-fs length was significantly associated with progression to KF (HR 1.19 per 100 amino acids, 95% CI 1.03–1.37, p = 0.015), whereas total protein length was not (HR 1.07, 95% CI 0.95–1.21, p = 0.28). Adjustment for sex did not modify this correlation.

Longer proteins were also associated with a steeper eGFR decline (Spearman rho = −0.37, p = 0.009 for total protein length; rho = −0.36, p = 0.010 for MUC1-fs length; **Figure 2A, 2B**). Median slopes were steeper in the long-protein groups after dichotomisation at the median (−2.24 vs −2.00 mL/min/1.73 m^2^/year, p = 0.033 for total protein length; −2.56 vs −1.75, p = 0.0085 for MUC1-fs length; **Figure 2C, 2D**). In linear mixed models with a random intercept for kindred, continuous measures for protein and MUC1-fs length did not correlate with eGFR slope (**Supplementary Table S3**), whereas the difference between MUC1-fs length groups remained significant (−0.72 mL/min/1.73 m^2^/year, 95% CI −1.32 to −0.12, p = 0.028; **Supplementary Table S3**).

**Figure 2.**
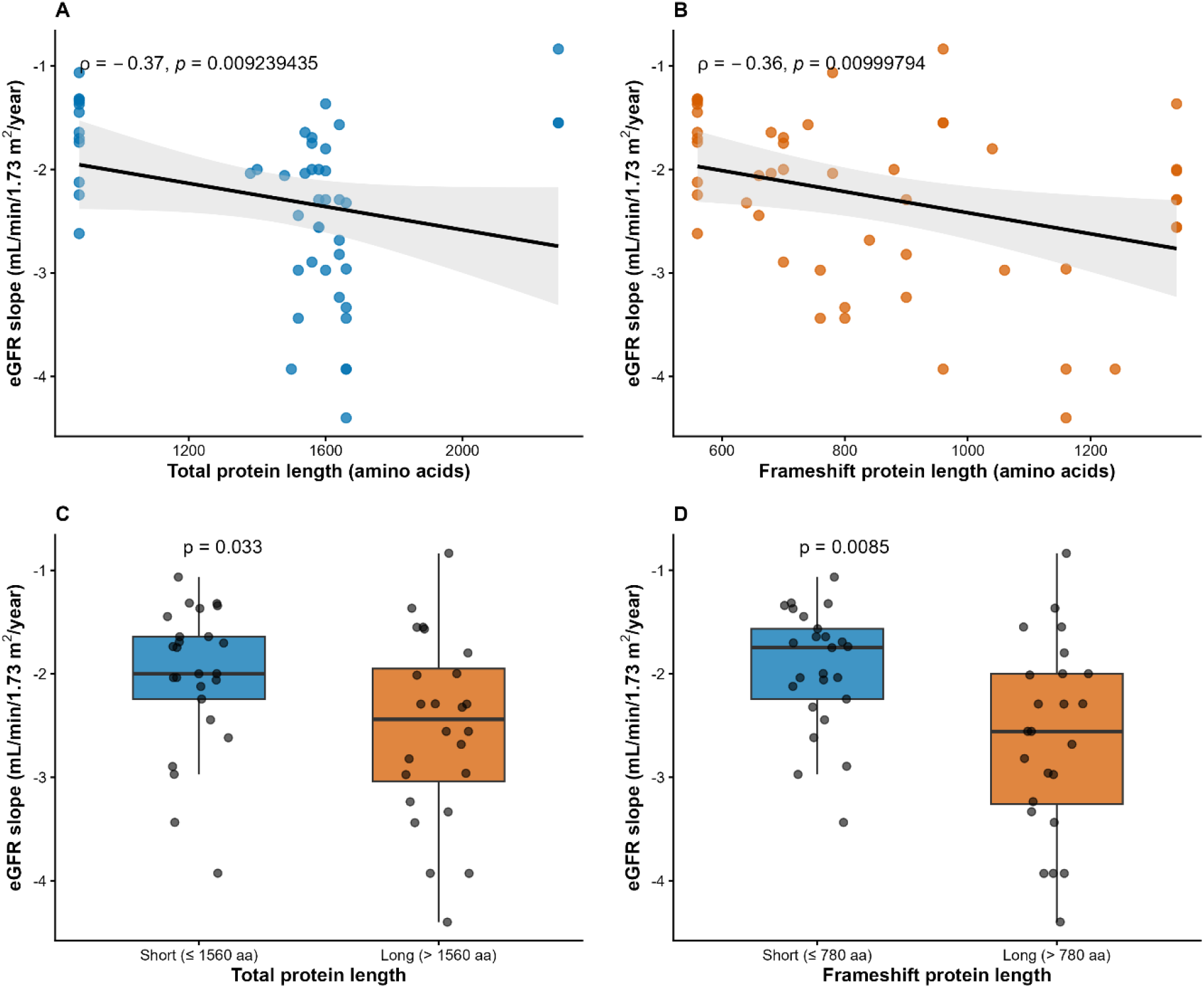
eGFR slope according to total and MUC1-fs lengths. Relationship between protein length and rate of renal function decline in the n = 49 patients with at least two eGFR measurements available. (**A, B**) Individual eGFR slopes plotted against total protein length (**A**) and MUC1-fs length (**B**), expressed in amino acids. Solid lines represent the least-squares regression line and shaded areas its 95% confidence interval; rho and the corresponding p-value are from Spearman rank correlation. (**C, D**) eGFR slopes in patients dichotomised at the median value of total protein length (**C**, 1560 amino acids) and of MUC1-fs (**D**, 780 amino acids). Boxes show the median and interquartile range, whiskers extend to 1.5 times the interquartile range, and individual patients are overlaid as jittered points; p-values are from the Wilcoxon rank-sum test. A more negative slope indicates faster loss of renal function. For patients who reached kidney failure, an eGFR of 10 mL/min/1.73 m^2^ was assigned at the date of dialysis initiation.

### Disentangling total from MUC1-fs protein length

We studied the association under five handlings of familial clustering (**Figure 3**). The Hazard Ratio (HR) for MUC1-fs length was stable throughout, between 1.19 and 1.20. In leave-one-kindred-out analyses, it ranged from 1.11 to 1.28 (median 1.19) and remained significant in 22 of 24 analyses, the exceptions being the kindreds at the extremes of MUC1-fs length. Sampling one individual per kindred over 200 draws gave a median HR of 1.20 (2.5th–97.5th percentiles 1.06–1.30). Total protein length remained close to the null throughout.

**Figure 3.**
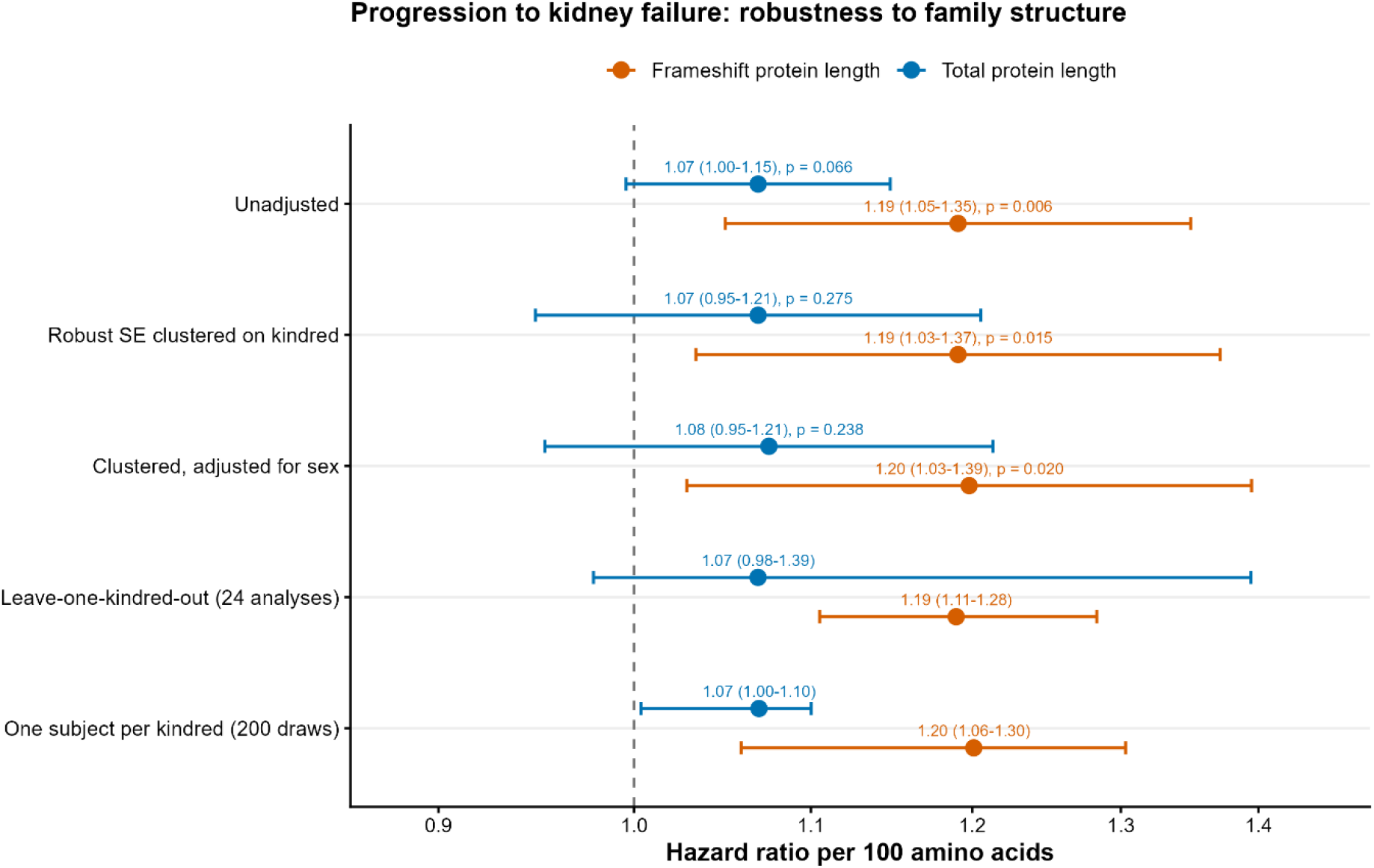
Hazard ratios for progression to kidney failure across five approaches to familial clustering. Hazard ratios (HR) for progression to kidney failure, per 100 amino acids, for MUC1-fs length (orange) and total protein length (blue), under five handlings of familial non-independence. Rows 1–3 are Cox models unadjusted, with robust standard errors clustered on family, and additionally adjusted for sex; with bars showing 95% confidence intervals. In the leave-one-family-out analysis, each of the 24 independent units (11 families, 13 singleton kindreds) was removed in turn; in the random-draw analysis, one individual per unit was sampled over 200 replicates. For these two rows the point is the median hazard ratio and the bar its range across analyses (full range and 2.5th–97.5th percentiles, respectively), not a confidence interval. The dashed line indicates HR = 1; the × axis is logarithmic. The estimate for MUC1-fs length remained above unity under all five specifications, whereas that for total protein length was closer to the null and its confidence interval included 1 in the clustered models

When combined into the same Cox model, the effect of total protein length fell to the null (HR 1.02, 95% CI 0.89–1.17), whereas MUC1-fs length retained a borderline association (HR 1.17, 0.98–1.39, p = 0.075), with low collinearity (VIF 1.37; **Supplementary Table S2**). Adding MUC1-fs length to a model already containing total protein length improved fit (p = 0.049), whereas the converse did not (p = 0.66), and MUC1-fs length gave the lower Akaike information criterion (**Supplementary Table S4**). A residualisation analysis was consistent but less conclusive.

Dividing patients into tertiles of MUC1-fs length (n = 20, 16 and 13) showed a progressive increase in risk: compared with the shortest tertile, the HR was 2.58 (95% CI 0.92–7.25, p = 0.073) in the intermediate tertile and 4.53 (95% CI 1.65–12.45, p = 0.003) in the longest (**Supplementary Figure S1**). A test for linear trend across tertiles was significant (HR 2.14 per tertile, 95% CI 1.30–3.54, p for trend = 0.003), indicating a dose-dependent relationship rather than an effect confined to a single threshold.

## Discussion

In 49 patients with ADTKD-MUC1, the length of the MUC1-fs protein predicted age at KF, with a graded effect across tertiles and an approximately fourfold difference in risk between extreme tertiles. Total protein length showed a weaker association that was attenuated once frameshift length was accounted for, indicating that the signal resides in the aberrant portion of the protein rather than in overall allele size.

While this study was ongoing, Vrbacká et al.^9^ reported, using SMRT sequencing in the Wake Forest ADTKD registry, that faster progressors carry more repeat units on the mutation-bearing allele, particularly more frameshifted repeats, explaining approximately 6% of the variance in progression^9^. Our findings are consistent with theirs despite differences in sequencing technology and extend their work by examining a time-to-event outcome and explicitly accounting for kinship, after which the association persisted.

This is biologically coherent. MUC1-fs is not secreted and instead accumulates in a TMED9-positive early-secretory compartment, where it is toxic to tubular cells. Clearing the fs-protein reverses the cellular phenotype^4^. A longer MUC1-fs plausibly imposes a greater burden per molecule, through enhanced aggregation, slower clearance or more extensive sequestration of the secretory machinery, consistent with the graded rather than threshold effect we observed.

The association was weaker for eGFR slope: neither continuous measure survived adjustment for kinship, though the MUC1-fs group difference persisted. Because genotype varied only between kindreds, its effect was difficult to disentangle from the kindred-level random intercept; notably, kindred membership alone accounted for 70% of the slope variance. Limited power and unmeasured factors (such as genetic background and environment) likely contribute to disease severity as well; disentangling them will require larger cohorts and possibly the use of polygenic scores.

Several limitations should be acknowledged. The cohort is small and, more importantly, comprises only 24 independent genotypes, which caps the information available on genotype– phenotype correlation. The data are retrospective, and eGFR slopes were derived from measurements collected in routine care with variable intervals. Finally, the mutant allele was characterised by long-read sequencing in 28 patients (24 unrelated patients and 4 affected family members) and inferred in the remaining 21 affected relatives.

In conclusion, the position of the frameshift within the *MUC1* VNTR determines the length of the resulting MUC1-fs protein, and this length predicts age at KF in a graded manner, robust to familial structure. ADTKD-*MUC1* should therefore no longer be regarded as a genotypically uniform disease. Replication in a larger multicentre cohort would help strengthen these findings and assess the value of reporting the position of the frameshift in order to refine prognosis.

## Data Availability

All data produced in the present study are available upon reasonable request to the authors

## Ethics and reporting

Clinical data were collected from patients’ medical records, at Necker-enfants malades University Hospital. Written informed consents were obtained. Regulatory and ethical approvals are detailed in the supplementary material. This study is reported in accordance with the STROBE statement for cohort studies.

## Authorship

GD supervised and conceptualized the study, MO performed formal analysis and first draft, MO, HS, AC, ZAA, MF, MMG, VM and OG performed the investigation HS, AC, BK, LH, CA and GD reviewed the draft. Final approval: MO, HS, AC, JK, ZAA, MF, MMG, VM, OG, BK, LH, CA and GD.

## Data Availability Statement

The data that support the findings of this study are available from the corresponding author, upon reasonable request.

## Acknowledgments

The authors gratefully acknowledge the patients and their family.

