## Supplementary Tables and Figures for "The frameshifted VNTR length determines the kidney survival in *ADTKD-MUC1*"

Affiliations:

^#^Collaborators: Tayeb Bensalem, Guillaume Canaud, Christophe Charasse, Christian Combe, Mohamed Said Dahmoune, Myriam Dao, Elsa Ferriere, Aurélie Hummel, Dominique Joly, Charlène Levi, Karine Moreau, Christiane Mousson, Marie-Noëlle Peraldi, Thierry Petitclerc, Romain Pszczolinski, Philippe Rémy, Philippe Vanhille, Vincent Vuiblet

**Keywords**: ADTKD-*MUC1*, genotype/phenotype correlation, Long-Read Sequencing Technology, genetics, kidney genetics.

**Supplementary :**

** Supplementary Table S1. Characteristics of the MUC1 cohort**

| **Characteristic** | **N = 49**^1^ |
| --- | --- |
| **Sex** |  |
| Female | 20 (41%) |
| Male | 29 (59%) |
| **eGFR slope (mL/min/1.73 m²/year)** | -2.1 (-2.8--1.6) |
| **Gout** |  |
| No | 20 (71%) |
| Yes | 8 (29%) |
| NC | 21 |
| **Renal cysts** |  |
| No | 13 (54%) |
| Yes | 11 (46%) |
| NC | 25 |
| **Total protein length (aa)** | 1,560.0 (1,380.0-1,640.0) |
| **MUC1-fs length (aa)** | 780.0 (660.0-1,040.0) |
| **Mutant allele repeats** | 78.0 (69.0-82.0) |
| **Mutation position in repeat unit** | 27.0 (16.0-43.0) |
| **Mutated repeat unit^‡^** |  |
| X | 27 (55%) |
| B | 11 (22%) |
| D | 9 (18%) |
| Others | 2 (4.0%) |
| ^1^n (%); Median (Q1-Q3); NC: non communicated; aa: amino acids; ‡: repeat unit as described by *Kirby et al. 2013*. | |

|  | **MUC1-fs length** | | | **Total protein length** | | | **Both measures combined** | | |
| --- | --- | --- | --- | --- | --- | --- | --- | --- | --- |
| **Characteristic** | **HR** | **95% CI** | **p-value** | **HR** | **95% CI** | **p-value** | **HR** | **95% CI** | **p-value** |
| Per 100 amino acids | 1.19 | 1.03, 1.37 | 0.015 |  |  |  |  |  |  |
| Per 100 amino acids |  |  |  | 1.07 | 0.95, 1.21 | 0.28 |  |  |  |
| Total (per 100 aa) |  |  |  |  |  |  | 1.02 | 0.89, 1.17 | 0.8 |
| Frameshift (per 100 aa) |  |  |  |  |  |  | 1.17 | 0.98, 1.39 | 0.075 |
| The proportional hazards assumption was satisfied for both measures (p = 0.82 and p = 0.77). Abbreviations: CI = Confidence Interval, HR = Hazard Ratio | | | | | | | | | |

** Supplementary Table S2. Cox proportional hazards models for kidney failure-free survival** (robust standard errors clustered on family)

*****Supplementary Table S3. Linear mixed models for eGFR slope, with a random intercept for kindred*****

| Analysis | Beta | 95% CI | P |
| --- | --- | --- | --- |
| Total protein length, per 100 aa | -0.07 | -0.18 to 0.04 | 0.216 |
| MUC1-fs length, per 100 aa | -0.12 | -0.26 to 0.01 | 0.097 |
| Total protein length, long vs short | -0.41 | -1.05 to 0.23 | 0.220 |
| MUC1-fs length, long vs short | -0.72 | -1.32 to -0.12 | 0.028 |
| Beta is the change in eGFR slope (mL/min/1.73 m2/year) per 100 additional amino acids, or between length groups dichotomised at the median. All models include a random intercept for kindred (24 independent units); p-values use Satterthwaite's approximation. Because genotype is constant within each kindred, its effect is entirely a between-kindred effect, and the random intercept competes with it for the same variance: Kindred membership alone accounted for 70% of the variance in eGFR slope. After accounting for MUC1-fs length, 68% of the remaining variance in eGFR slope lay between kindreds. | | | |

**Supplementary Table S4. Comparison of MUC1-fs and total protein length as predictors of kidney failure**

| Approach | MUC1-fs length | Total protein length |
| --- | --- | --- |
| Added to a model containing the other measure (chi2) | 3.880 | 0.190 |
| Added to a model containing the other measure (P) | 0.049 | 0.663 |
| Akaike information criterion | 190.000 | 193.700 |
| Residualised effect (P) | 0.103 | 0.575 |

Model comparison rests on the partial likelihood, which the specification of robust standard errors leaves unchanged; the likelihood ratio tests and information criteria below were therefore computed on unclustered models. They supported a consistent hierarchy: adding MUC1-fs length to a model already containing total protein length improved fit (p = 0.049), whereas adding total protein length to a model already containing MUC1-fs length did not (p = 0.66), and MUC1-fs length gave the lower Akaike information criterion. The residualisation analysis was consistent but less conclusive: neither residualised component reached significance, although the effect of MUC1-fs length independent of total protein length was closer to the conventional threshold (p = 0.10) than the converse (p = 0.58). Residualised models, like all estimates reported in the main text, use robust standard errors clustered on kindred.

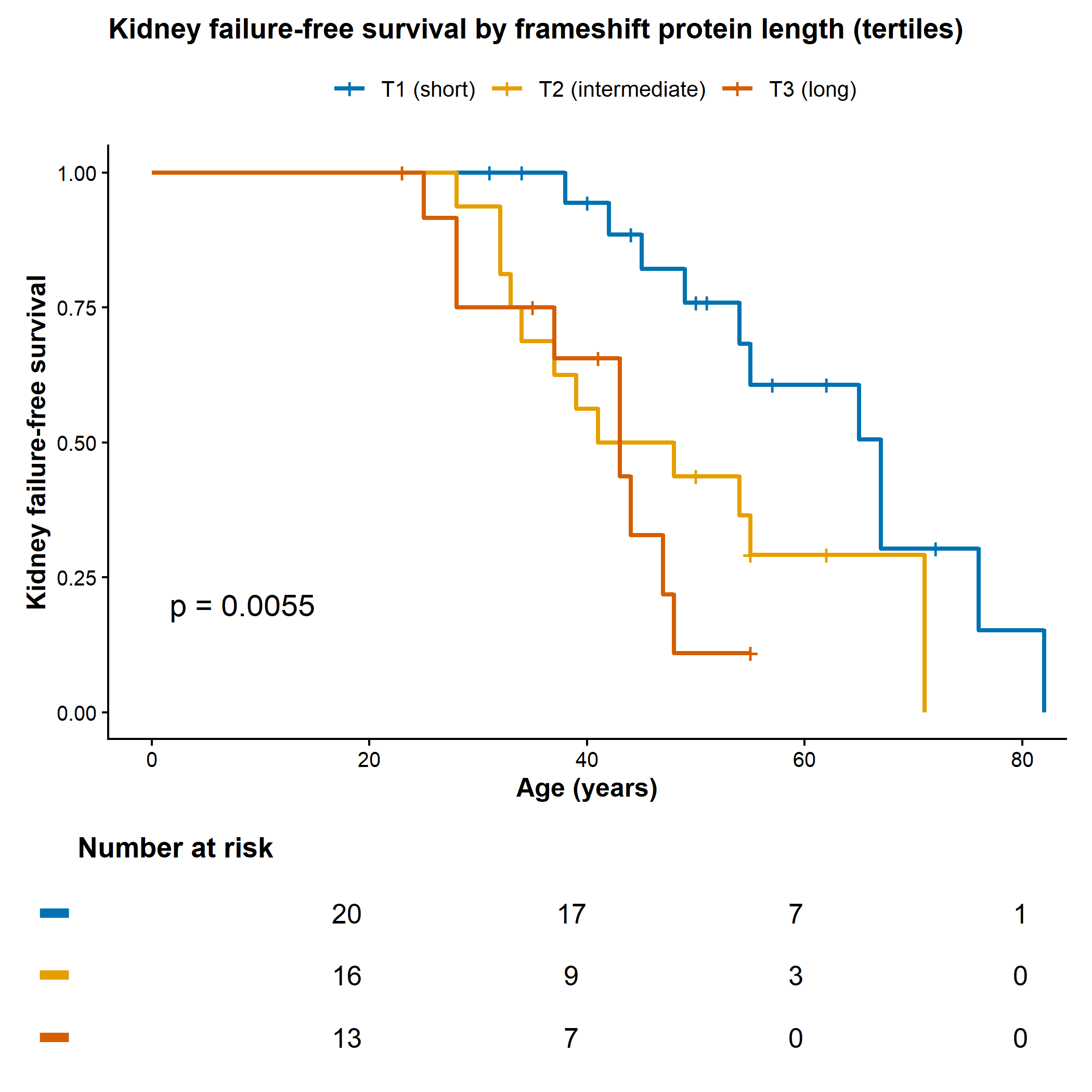

**Supplementary Figure S1.** **Kidney failure-free survival according to tertiles of MUC1-fs length.** Kaplan-Meier estimates of kidney failure-free survival, with age as the time scale, in the 49 patients grouped by tertile of MUC1-fs length (T1, 560–700 aa, n = 20; T2, 701–960 aa, n = 16; T3, 961–1340 aa, n = 13). Vertical ticks denote censored observations and numbers at risk are shown below the panel. Survival differed significantly across the three groups (p = 0.0055, global log-rank test). In a Cox model with robust standard errors clustered on family, each increasing tertile was associated with a hazard ratio of 2.14 (95% CI 1.30–3.54, p = 0.003; test for trend). This analysis is presented as a sensitivity analysis to the dichotomised model of **Figure 1B** and shows a graded rather than threshold relationship between MUC1-fs length and kidney failure.

**Supplementary material:**

Supplementary material will be made available upon publication.
